# Recessive and sex-dependent genetic effects in primary hypertension

**DOI:** 10.1101/2022.05.31.22275828

**Authors:** Roei Zucker, Michal Linial

## Abstract

**Background:** Essential hypertension is a polygenic disease that affects almost half of the adult population in the USA. It is a major risk factor for renal, cerebrovascular, and cardiovascular diseases. Previous studies used UK-Biobank (UKB) GWAS results for hypertension to create a polygenic risk score (PRS), with the top and bottom 5% of the PRS translating to a 4-fold difference in the estimated risk. The heritability of hypertension is estimated to be high (30–60%), yet the underlying mechanisms and the associated genes are largely unknown.

**Methods:** In this study, we used a gene-based method, the proteome-wide association study (PWAS), to detect associations mediated by the effects of variants on protein function. PWAS was applied to individuals of European ancestry from the UKB, with 74,090 cases of clinical diagnosis of essential (primary) hypertension (ICD-10, I10) and 200,734 controls. PWAS aggregates the signal from all variants affecting each coding gene and provides scores for dominant, recessive, and hybrid genetic heritability.

**Results:** PWAS identified 70 statistically significant associated genes (FDR-q-value <0.05) and 127 genes with a weaker threshold (FDR-q-value <0.1). The overlap with GWAS summary statistics (total 1,362 genes) is only partial, with 23 and 62 genes identified exclusively by PWAS from a total of 70 and 127 genes, respectively), among them 18% were assigned recessive inheritance. Furthermore, PWAS analysis, separately performed on females and males from UKB genotyping imputed data, revealed sex-dependent genetics. There are 22 genes unique in females, with only 2 in males. We identified 6 female-specific genes that were not identified by PWAS for the entire group (70 genes). Only one associated gene (SH2B3) is shared between the sexes. Many of the female-significant genes from PWAS are enriched in cellular immunity functions.

**Conclusions:** We conclude that hypertension displays sex-dependent genetics with an overlooked recessive inheritance, postulating that the underlying mechanism is substantially different for males and females. Studying hypertension by a gene-based association method improves interpretability and clinical utility.

## INTRODUCTION

Essential hypertension is a common medical condition among the adult population, and its prevalence continues to rise. It is estimated to affect over 1.2 billion people globally. However, only 42% of the adults with hypertension are diagnosed and treated. The diagnosis is based on persistent high blood pressure (BP). The cause of BP is mostly unknown, but many studies have confirmed its complex physiology including marked genetic component [1-3]. In the USA, almost half of the population >20 years old meets the clinical criteria, with a ratio of 1.2 of affected males relative to females [4, 5]. While the difference in occurrences implies a sex difference, the etiology is unknown [6]. Hypertension increases the risk for chronic kidney disease, cerebrovascular disease, and cardiovascular disease (CVD), including stroke [7 47].

According to family and twin studies, hypertension heritability account for 30–50% of the variance in BP. Attempts to identify the genetic basis for hypertension in affected families have resulted in a long list of variants [8]. Genotyping data from large population studies revealed an SNP-heritability of 0.2632, putting it in the top 5% of heritable phenotypes studied [9]. The analysis of rare monogenic syndromes of hypertension highlighted pathways involving sodium handling and steroid hormone metabolism, including mineralocorticoid receptor activity [2]. While several regulatory networks have been proposed to contribute to hypertension and BP based on animal models [3, 10], the mechanistic understanding remains obscure [11]. To better understand hypertension, it appears that environmental effects and gene interactions will need to be considered [12].

GWAS experiments are carried out to identify common candidate genetic variants in the studied population [13]. Despite the clear advances and simplicity of the GWAS protocol [14], it suffers from poor biological interpretation as many of the identified variants are localized in introns, intergenic regions, and unannotated genomic segments. The small effect size attributable to common variants also limits the interpretation of the genetic basis for most polygenic diseases, including hypertension [15]. An increase in the GWAS statistical power was achieved by using summary statistics of correlated traits [16]. There have been numerous GWAS that sought to identify variants attributable to BP [17]. Early attempts to identify associated variants from GWAS for hypertension [3, 18] identified only 43 genetic variants, with a single gene (ATP2B1) uniquely associated with hypertension [18]. A meta-analysis combining several GWAS for BP (systolic, diastolic, pulse pressure) performed on European ancestry [19] and on the UK-Biobank (UKB) identified 107 replicated loci which were assigned to 212 candidate genes (∼330k participants) [7]. However, only a few of the informative SNPs affect the coding of the candidate gene. The number of loci was expanded as the number of genotyped participants increased. From the Million Veteran Program (MVP) and the UK-Biobank (UKB), covering 460k participants (total 840 associated genes), a set of 208 novel common variants for BP were reported. These studies suggested that significant effects of the identified SNPs act on gene regulation (e.g., enhancers, promoters, and miRNA binding sites) [20]. Based on compilation and analysis of over 1 million people of European ancestry, >500 novel BP loci have been identified [21, 22]. Many identified variants fail to comply with the statistical threshold for a valid association, but are useful for developing a phenotype-based polygenic risk score (PRS). The PRS approach was developed for prediction purposes where a disease risk associated with an individual’s genetics is tested with respect to the population cumulative data. In the case of hypertension, even the best PRS provides only a small fraction of the variation in the population. The top and bottom 2.5% of the PRS had 2-fold higher and lower risks of hypertension, respectively, when compared to the general population [23]. At present, PRS for hypertension has limited clinical usability [24].

In this study, we applied a gene-based method from the Proteome-wide association study (PWAS) [25] that detects gene-phenotype associations through the effect of variants on protein function. PWAS aggregates the signal from all variants affecting each protein-coding gene with the assumption of dominant and recessive inheritance. While routine applications of GWAS assume additive heritability for genetic effects, ignoring alternative heritability modes [17], recessive effects account for a substantial genetic signal in complex traits (e.g., cancer) [26]. We identified a gene-association predisposition for hypertension that is sex-dependent. We further propose that genetic signals in females include an overlooked immunological-related signal. These findings, and the gene-based PRS presented for hypertension, can improve our understanding of the biology of hypertension and eventually improve disease management.

## METHODS

### UKB processing

The UK Biobank (UKB) is a population-based database with detailed medical, genotyping, and lifestyle information covering 500k people at ages 40-69 across the UK who were recruited from 2006-2010. The analyses were based on the 2019 UKB release unless otherwise mentioned. We restricted the analysis to self-reporting of British, Irish, or other Caucasian background [codes 1, 1001, 1002, 1003, respectively, in Ethnic background, UKB data field 21000]) and classified as Caucasians based on their genetic ancestry (Genetic ethnic group, data-field 22006). We further removed genetic relatives, by randomly keeping only one representative of each kinship group.

Disease classification is based on clinical information encoded by ICD-10 codes of hypertension (I10) within main or secondary diagnosis codes (UKB data fields 41202 and 41204, respectively). In cases where multiple values were reported for a specific field (in a continuous phenotype) the maximal value is considered. The filtered UKB included 154,588 males, 178,827 females. Among them 74,090 were diagnosed by I10, with 40,358 males and 33,732 females (45.5%).

### Hypertension genetic analysis

The UKB released genotyped data for all participants. The UKB genotyping scheme is based on ∼820,000 preselected genetic variations (from genotyping data of the UKB Axiom Array). For hypertension (I10), we tested 804,069 informative markers. Based on UKB imputation protocol, the number of variants was expanded to the 97,013,422 imputations variants provided by the UKB (with about 9 M variants that passed quality control). In hypertension, we analyzed the variants positioned at the coding regions (for PWAS, and the matched GWAS). All together we tested all variants that are included in the 18,053 coding genes, including splicing variants. This set included 639,323 of the 97,013,422 of the whole genome imputed variants.

### Comparative analysis

We used the Open Targets platform to select current knowledge on hypertension. Open Targets [27] (OT, dated 3/2022) compiled a list of associated genes from large scale GWAS resources. It is a public database that unifies evidence for drugs, their targets, and their associations with human diseases. From the OT database, the collection of genetic associations for hypertension includes 1,362 genes (out of 5,328 genes with some evidence) that were ranked based on GWAS summary statistics [28]. We extracted the SNPs associated with hypertension as reported by OT and used OT genetic association scores to list the top variants associated with hypertension for comparative analysis. The OT compiled information on essential hypertension from 5 studies with summary statistics. Most informative associations were reported by SAIGE_UKB (2018, 131 leading variants), FINNGEN_R5 (2021, 35 loci) and 19 independent loci [29]. All these studies were compiled to a unified gene list by OT genetic-association normalized scores (ranges 0-1.0). The gene set and their statistics are listed in Supplementary **Table S1**.

### PWAS methodology

PWAS was developed as a gene-based association method and was shown to complement GWAS for a large collection of human diseases and human traits [25]. In a nutshell, PWAS methodology assumes that causal variants in coding regions affect phenotypes by altering the biochemical functions of the encoded protein of a gene. The functional impact rating at the molecular level (called FIRM) pretrained machine-learning (ML) model, called FIRM, is then used to estimate the extent of the damage caused to each protein in the entire proteome [30]. FIRM used 1109 numerical features which are used by the ML model to predict variant-centric effect score (https://github.com/nadavbra/firm). FIRM performance was tested with respect to 3-fold cross validation on ClinVar with AUC = 90% (precision = 86%, recall = 85.5%) and specificity = 78.4%, F1 = 85.8% and accuracy = 82.7% [30]. Per-variant damage predictions are then aggregated at the gene level, in view of each sample’s genotyping, generating a protein function effect score for dominant or recessive effects on phenotypes (and a model for either mode of inheritance, called hybrid model). In addition, PWAS provides a generalized model that uses both the dominant and recessive values. The gene-based aggregation of the FIRM score associated with each variant allows the result of PWAS to cumulatively enhance weak signals associated with individual SNPs. On average, each gene is affected by 35.4 nonsense and missense mutations.

### Comparable GWAS analysis

To perform an unbiased comparison between PWAS and GWAS for hypertension, we used the same list of 17 covariates composed of sex (binary), year of birth (numeric), 40 principal components of the genetic data that capture the ancestry stratification were provided by the UKB (numeric), the UKB genotyping batch (one-hot-encoding, 105 categories), and the UKB assessment centers associated with each sample (binary, 25 categories). We tested the 804,069 genetic markers genotyped by the UKB (raw genetic markers) and the imputed coding variants that included 639,323 of the 97,013,422 imputated variants provided by the UKB. We tested the imputed variants that appeared to affect the coding region of 18,053 protein-coding genes. These are the identical set of variants (total 639,323) that PWAS considered. We made sure that PWAS and GWAS have access to data of the same variants. When testing raw genetic markers with GWAS, we used the convention of 0,1,2 variant encoding to specify the number of the alternative allele. For the imputed variants we calculated the probabilistic expected number of alternative alleles [25].

### Functional effect scores

In PWAS, each non-synonymous variant is assigned a functional effect score that aims to capture its propensity to damage the gene’s protein product, considering missense, nonsense, frameshift, in-frame indel, and canonical splice-site variants. The predicted effect score of a variant is a number between 0 (complete loss of function) and 1 (no functional effect). Intuitively, it reflects the probability that the affected gene retains its function given the variant. The FIRM captures gene function in its broadest sense allowing PWAS to model protein-phenotype effect of hypertension. Variations other than non-synonymous (i.e., nonsense, frameshift, splice site variants) scored 0 for a complete loss of function. In-frame indels were assigned an effect score based on some variants rule-based formulas [25]

### Statistical tests

For the effect size of a gene or variant, we applied the statistical measure of the Cohen’s d value to measure the difference between two means divided by a standard deviation (SD) for the data, also known as standardized mean difference. Formally, Cohen’s d = (M2 - M1) ⁄ SD_pooled_; where SD_pooled_ = √((SD_1_^2^ + SD_2_^2^) ⁄ 2)). The directionality of the effect is determined by defining M1 and M2 as cases and controls, respectively. Importantly, the effect-size metric results in PWAS and GWAS have opposite interpretations. In GWAS, a positive value indicates a positive correlation between the phenotype (e.g., hypertension) to the number of alternative alleles, thereby indicating a risk variant. In PWAS, on the other hand, positive values indicate a positive correlation with the gene effect scores, whose higher values mean less functional damage, thereby indicating protective genes. Thus, negative values are indicative of protective variants in GWAS vs. risk genes in PWAS.

## RESULTS

### GWAS results for ICD-10 for hypertension

Several GWAS on hypertension yielded over 1100 associated genes (Supplementary **Table S1**). However, the statistical power of these results is sensitive to the size of the cohorts, the exact definition of the phenotype (i.e. the ICD-10 diagnosis of I10 vs. self-reported hypertension). In addition, population structure, and the actual GWAS implementation impact GWAS results (Supplementary **Figure S1**). First, we performed GWAS for ICD-10 diagnosis of I10 (essential (primary) hypertension) of European origin from UKB. We considered a rich collection of 172 covariates (see Methods) for the 804,069 markers SNPs and identified 335 SNPs with an effective p-value <5e-07, with 205 of them complying with the widely accepted whole genome threshold (p-value <5e-08; Supplementary **Table S2)**. We considered two correlated measures for each significant GWAS variant, the p-value and Cohen’s d value. We calculated the directionality of the effect by calculating Cohen’s d value per variant (**Figure 1A)**. When negative, it indicates that cases (i.e., those diagnosed with I10) have a lower GWAS mean effect score compared to controls, meaning association with a reduced risk for hypertension. Conversely, a positive Cohen’s d indicates a risk. We show that the majority of the variants have a negligible effect size. Thus, we focused on 43 variants that are very significant (p-value <1e-12). Among them we considered Cohen’s d value larger than an absolute value of 0.03. There are 20 and 23 such variants with negative and positive Cohen’s d, respectively (**Table 1)**.

**Table 1.**
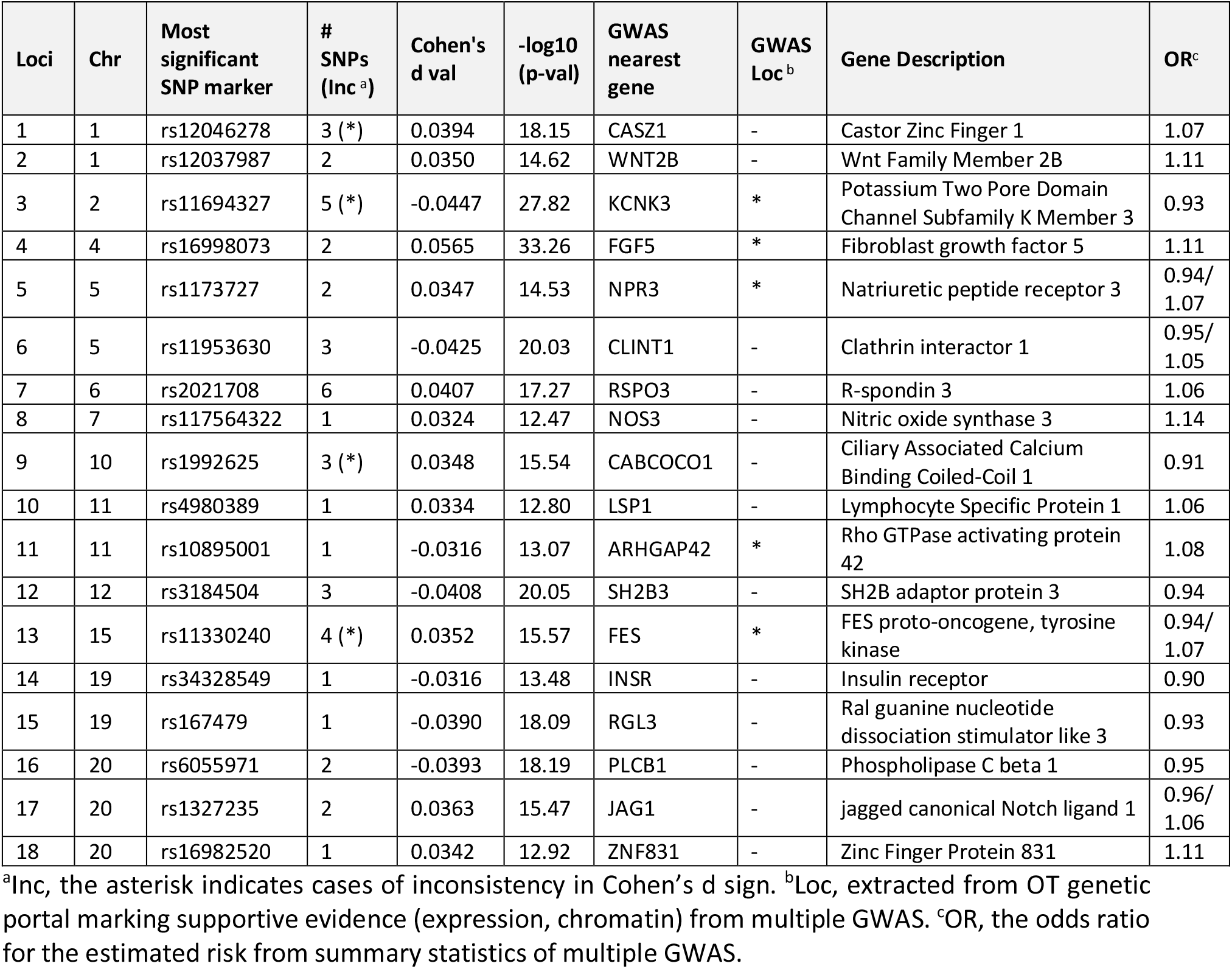
Independent loci derived from the top GWAS significant variants (804,069 markers).

**Figure 1.**
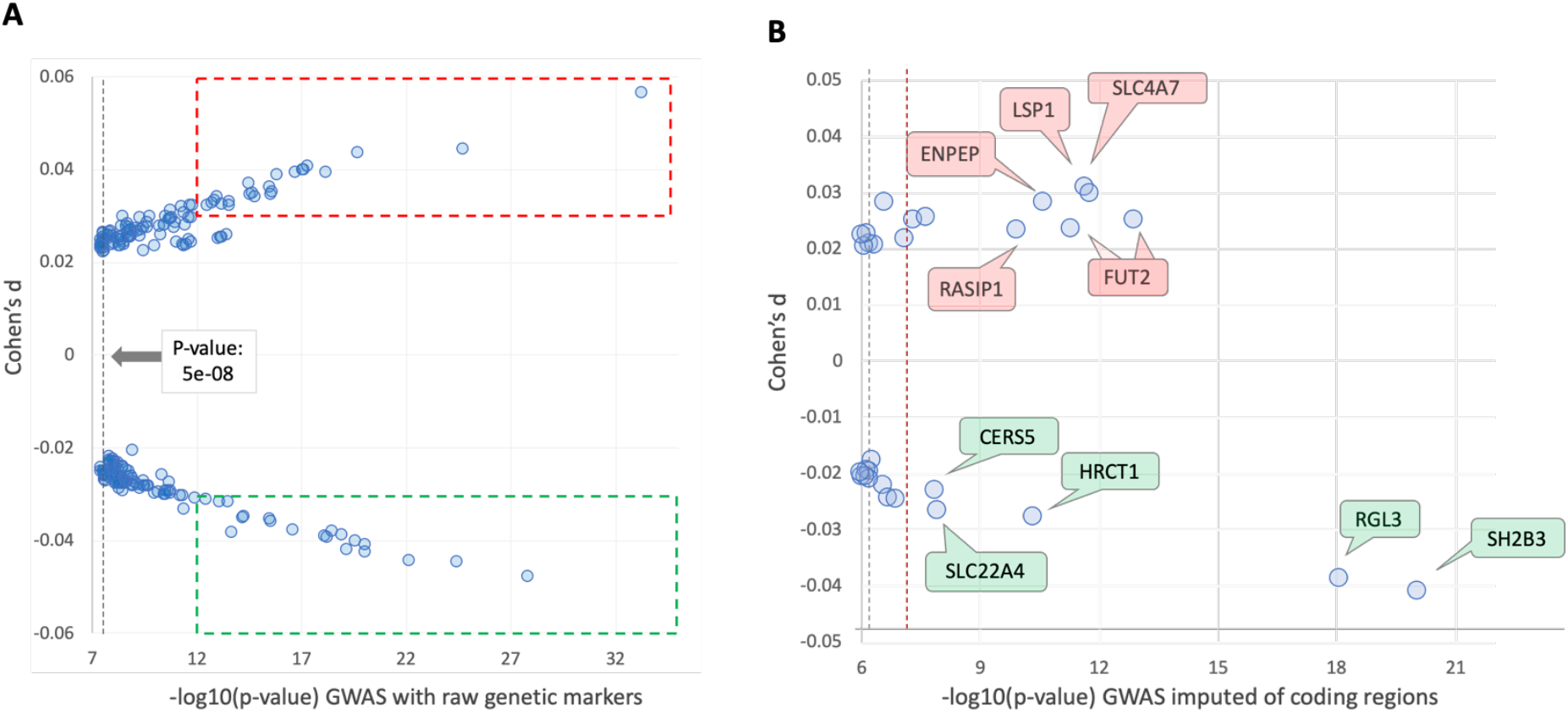
GWAS results for ICD-10 I10 (hypertension). **(A)** GWAS results for the UKB SNP markers (804,069) associated with Europeans with 205 significant variants (p-value <5e-08). Dashed frames show significant variants (p-value <1e-12) with a relatively high Cohen’s d value, larger than an absolute value of 0.03, with 20 and 23 variants with negative (green frame) and positive (red frame) Cohen’s d values, respectively. All 205 significant variants with their statistical analysis are listed in Supplementary **Table S2. (B)** GWAS-imputed variants of coding genes. Only 30 variants with a p-value <1e-06 are shown. The gray and red dashed lines mark the p-values <5e-7 and <5e-08 thresholds. The calculated Cohen’s d values are shown for the effect size of each variant within the coding region of a gene. Cohen’s d effect size values are marked in green and red frames around the associated genes to represent protective and risk variants, respectively. The SH2B3 gene has the highest confidence (p-value = 8.63E-21) and the strongest effect size (Cohen’s d value = -0.0408). SH2B3 variants were implicated in reducing the risk of hypertension (green). All 30 significant variants with their statistical analysis are listed in Supplementary **Table S3**).

**Table 1** summarizes current knowledge on these most significant variants and the candidate genes associated with them. All 43 variants (framed in **Figure 1A**) were assigned to 18 loci, with 12 loci supported by at least 2 significant variants **(Table 1)**. Interestingly, a third of the multiple variant loci showed some degree of directionality inconsistency in their effect size. The associated genes and calculated odds ratio (OR) for each variant were compared to that from OT reports obtained from the summary statistics from multiple resources. OR were compiled from summary statistics (see Methods), which are UKB based. For example, essential hypertension (non-cancer illness code), self-reported, with n= 361,141 including 93,560 participants as cases (OT, Neale v2 GWAS) and UKB SAIGE with n = 408,343 (77,977 case) [27]. Supporting evidence for the listed gene in the loci applied for only 5 genes (**Table 1**, asterisk in gene localization). Evidence for gene localization derived from the orthogonal external observations of eQTL, tissue expression, chromatin states and more.

The positions of most GWAS leading variants identified in **Figure 1A** are outside of gene coding regions and mostly occur in introns and intergenic regions. To extend the list of candidate-associated variants for hypertension while increasing the interpretability of GWAS analysis, we expanded the variant list to the imputed data assigned for the coding region of all 18,053 protein-coding genes. This included 639,323 of the 97,013,422 UKB imputed variants (see Materials and Methods).

**Figure 1B** shows Cohen’s d values of all variants that met a threshold of p-value <1e-06, with the 30 most significant coding region GWAS (total of 639,323 analyzed variants), among which 19 variants met the accepted exome-based threshold of p-value< 5e-07 (**Figure 1B**, red vertical dashed line). Among the genes with the strongest association and lowest Cohen’s d value are SH2B3 and RGL3, both of which are associated with protective variants for hypertension. Both are signified with high confidence (p-values) and relatively strong effect size (Cohen’s d values) (**Figure 1B**). We concluded that in the two settings of GWAS for essential (primary) hypertension (i.e., whole genome using genotyping and imputed set of coding regions, **Figure 1A** and **Figure 1B**, respectively), biological interpretation is lacking and the directionality of the effect of the variant on the associated gene (i.e., increasing or decreasing the hypertension risk) is often inconclusive.

### PWAS results for ICD-10 for hypertension

To overcome these inherently limitations of GWAS, we applied PWAS that exclusively focuses on the alteration in the coding region of the gene and assesses the impact of damaging SNVs on the protein function [25]. On average, for UKB genotyping data, each gene is affected by 35.8 nonsense and missense mutations (with a median of 26 SNPs per gene). For the ICD-10 I10 analysis (UKB 2019), we identified 70 PWAS genes (with an FDR q-value <0.05). The PWAS with an FDR q-value <0.1 yielded a list of 127 genes. These sets are referred to as PWAS (q-0.05) and PWAS (q-0.1) gene sets, respectively (Supplementary **Table S4**). **Figure 2A** compares the OT GWAS associated gene list (with 1362 genetic-associated genes, Supplementary **Table S1**) with the PWAS results. We identified 23 and 62 genes that were identified exclusively in the PWAS gene sets. OT associates each of the genes with a normalized score based on the genetic evidence. We observed that the 20% top scoring genes reported by OT match 45% of the 48 shared genes, confirming the enrichment in GWAS top-scoring genes and the PWAS list (**Figure 2B**).

**Figure 2.**
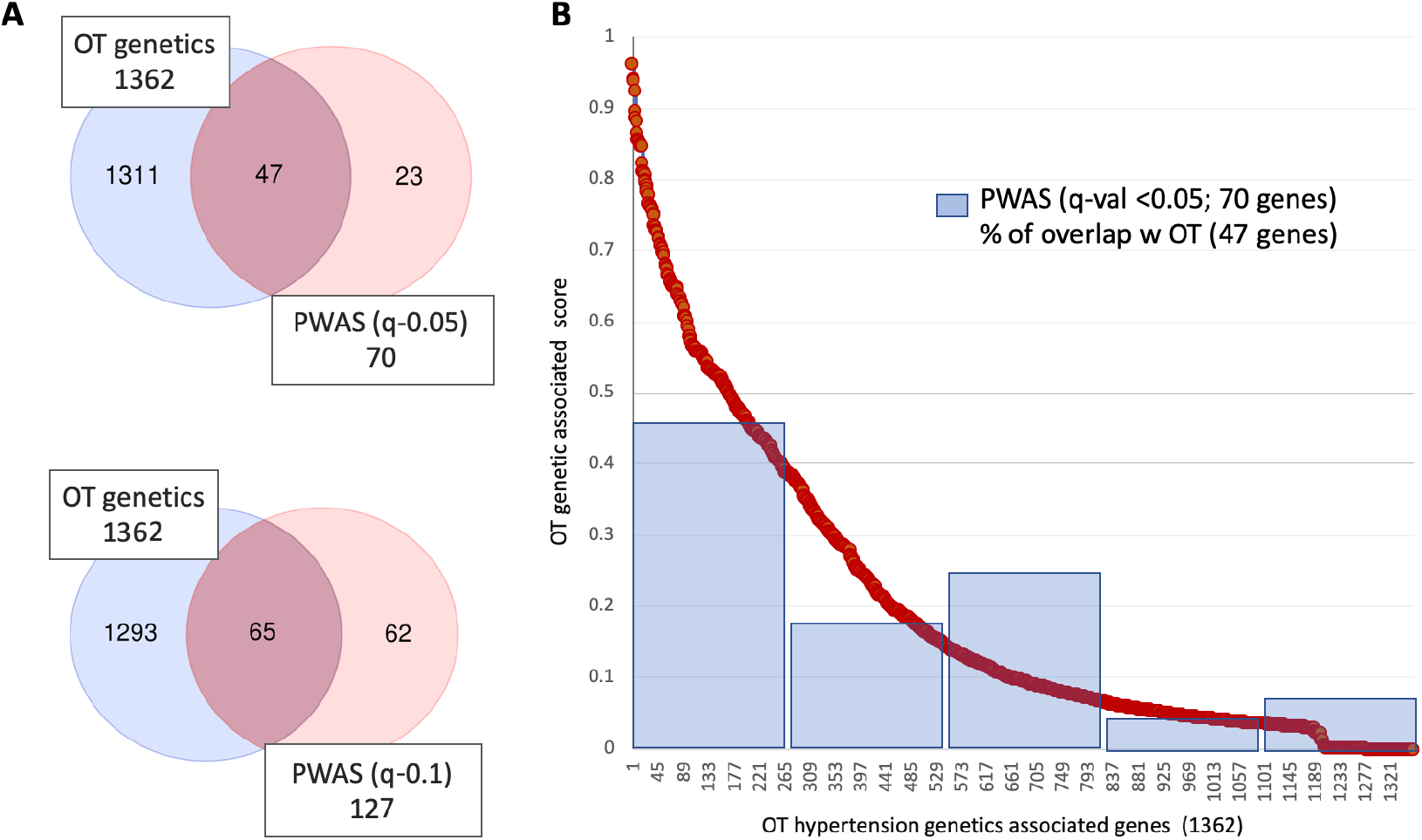
**(A)** Comparing the Open Targets (OT) GWAS associated gene list (total 1362 genes) and the PWAS results for q-value <0.05 and <0.1 with 70 and 127 genes, respectively. **(B)** OT genetic association scoring (in red) and the partition of the identified GWAS-PWAS shared 47 genes from the results of PWAS (q-0.05).

### Recessive effects are prevalent among PWAS significant genes

The PWAS aggregated score considers either dominant or recessive effects on phenotypes. In addition, PWAS provides a hybrid score that considers dominant and recessive inheritance modes. **Figure 3A** summarizes the partition of the 70 and 127 genes identified by PWAS (q-0.05 and q-0.1, respectively) for hypertension (ICD-10, I10) by their heritability results (Supplemental **Table S4**). The minimal value for assigning a gene with its preferred heritability model was set to -log10(p-value) <4. Among the genes identified by PWAS (q-0.05), only three out of 12 genes were assigned as recessively inherited genes and were not shared with the dominantly inherited gene group, while nine genes were shared by all three models (**Figure 3A**, top). These nine genes include ZNF493, SH2B3, SLC4A7, FUT2, REXO1, SLC22A4, FGF21, CYP11B2 and SWAP70 (Supplemental **Table S4**). The genomics of the CYP11B2 A similar analysis for the PWAS q-0.1 gene set of 127 genes showed that 35% of the recessive gene set was not shared by the dominant gene set (**Figure 3A**, bottom).

**Figure 3.**
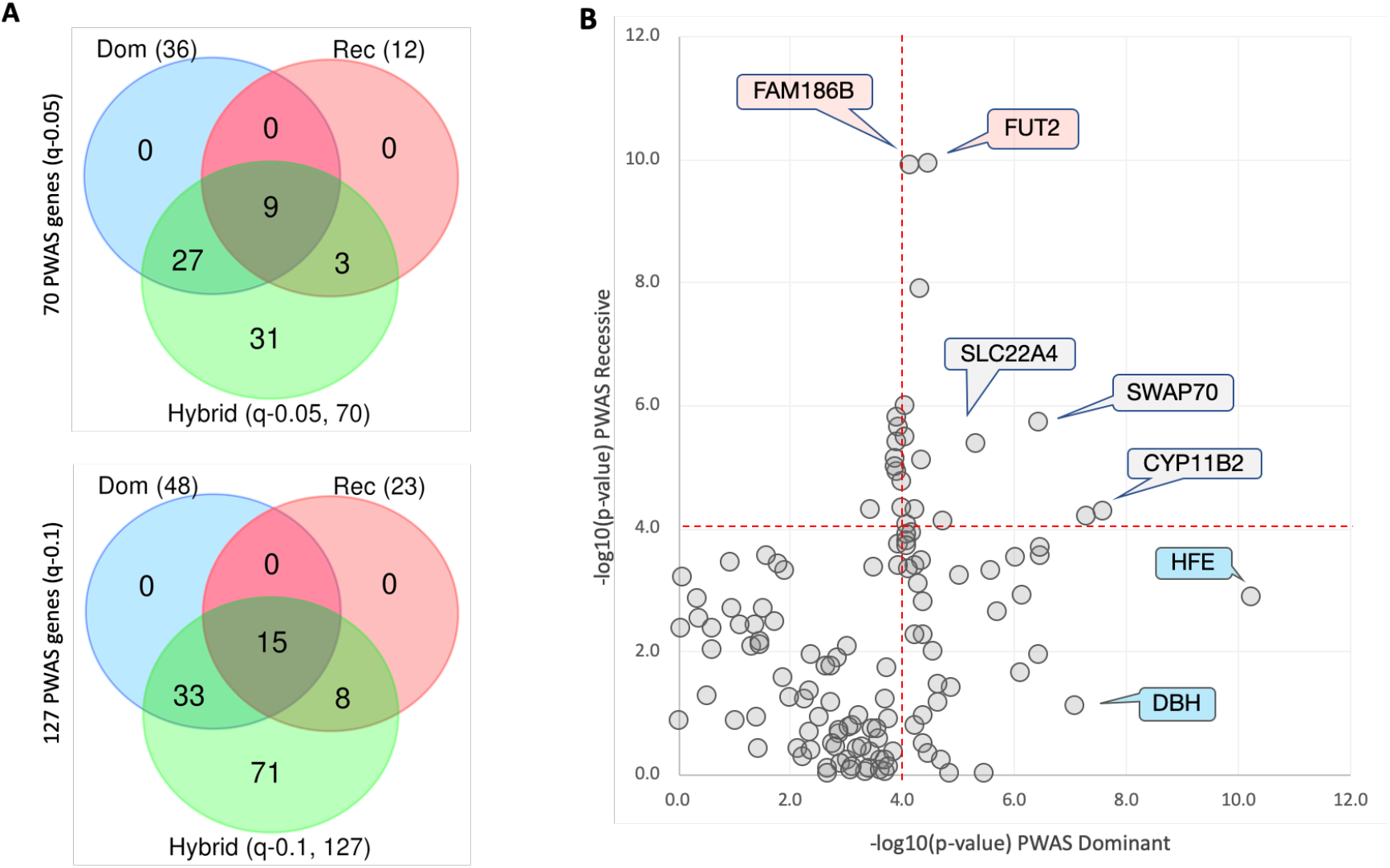
PWAS genes for ICD-10 hypertension (I10) according to three heritability models. **(A)** Venn diagram of PWAS identified genes by hybrid models and partitioned to dominant and recessive heritability with gene p-values <1e-04. Top: PWAS q-0.05 and Bottom: PWAS q-0.1. The number of each gene set is marked. Dom: dominant heritability genes; Rec: recessive heritability genes. **(B)** Scatter plot of gene effects for 127 genes associated with hypertension based on PWAS q-values <0.1. Each gene is indicated by -log10(p-value). Several representative genes are labelled as dominant (light blue) and recessive (line red). The most significant gene, SH2B3, with a -log10 (p-value) of 20.3 (dominant) and 8.8 (recessive) is not shown for keeping the scale of the axes. The dashed red lines show a threshold of -log10(p-value)>4 for Rec and Dom.

**Figure 3B** shows a scatter plot of -log10(p-value) for the dominant and recessive genes in the PWAS (q-0.1) gene set. Only a few genes were associated with stronger significance p-value of recessive relative to dominant assigned genes. Altogether, for 11 genes, the difference in the statistical p-values of dominant and recessive were >1e04, among them only two genes showed a more significant for the recessive vs. dominant p-values. In general, many of the genes identified by the hybrid models are also identified in the dominant mode. Note that a substantial number of genes (44% and 56% of PWAS q-0.05 and q-0.1, respectively) does not match either the dominant or the recessive model, but are still significant in the hybrid mode (i.e., model accounting for both dominant and recessive heritability).

We further analyzed the PWAS associated genes according to their gene effect size as tested by Cohen’s d, with negative and positive values being associated with increased and decreased risk, respectively (note the inverse interpretability for GWAS and PWAS, see Materials and Methods). We found that the effect size of 124 or 127 genes listed by PWAS (q-0.1) is larger than an absolute value of 0.03 (Supplementary **Table S4**). SH2B3 with the highest significant p-value (FDR q-value 9.8e-18) displays an effect size of > 0.038 and is indicative of a protective gene (Supplementary **Figure S2**). The other protective genes are EIF2S3 and PRKX (Cohen’s d values of 0.07 and 0.058, respectively; Supplementary **Figure S2**). At the PWAS gene level, 98% of the associated genes are of low effect size, supporting the polygenic nature of hypertension.

### GWAS significant variants of hypertension are sex dependent

We asked whether we could identify sex dependent genetic effects from the coding genes by performing GWAS analysis on 154,588 males and 178,827 females, 40,358 males and 33,732 females of whom were diagnosed with I10 (total of 74,090 cases). The calculated probability confirms a sex dependent hypertension diagnosis in our cohort (p-value = 1.9e-07). We analyzed the 18,053 protein-coding genes (and consensus splicing sites) with the 639,323 imputed variants (see Materials and Methods), where each gene has on average 35.8 variants. We collected 377 variants with a minimal - log10>3.0 in only the males or females. We avoided cases of both sex groups being significant to highlight variants that are characterized as being different in their significance by sex (Supplementary **Table S6)**

We then sought to identify variants with a marked difference between the sexes (Δz-values of >4.0). There are 63 and 43 such variants with positive and negative values, respectively (**Figure 4C**). The positive values (z-values ranging from 4.0 to 6.9,) are cases in which the z-value of a variant in females is positive while the same variant in males is of a negative directionality (**Figure 4C**, top panel). The negative values (z-values ranging from -4.0 to -6.0) are cases in which the opposite is true (**Figure 4C**, bottom panel). Inspecting this significant variant list (total 106) confirmed that the difference between the sexes is attributed to an invert directionality of the z-values for males and females (Supplementary **Table S5)**.

**Figure 4.**
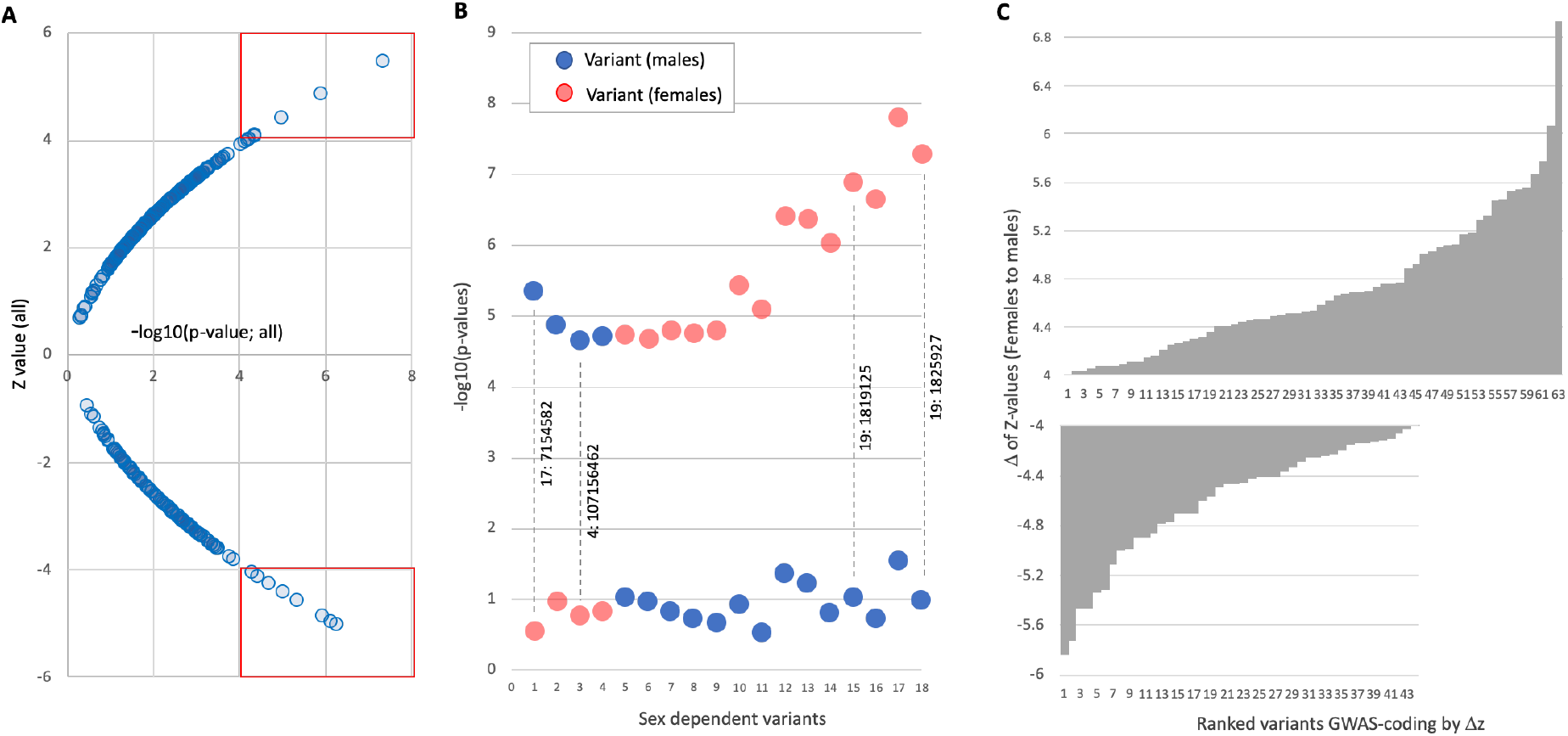
The results of GWAS performed on coding genes. **(A)** Effect size (z-score) for GWAS. The variants within the red frames in (A) are associated with at least -log10(p-value) >4. **(B)** Zoom-in on the 18 variants based on their -log10(p-value) for males and females (blue and red, respectively). For demonstration, we labelled four variants by their chromosomal location. **(C)** Sorted list of variants by the difference in Δz-values of females to males. There are 63 and 43 variants that are listed with a sex-dependent difference with Δz with absolute values of >4. In almost all variants, the -log10(p-value) of variants identified for females and males pointed in the opposite direction, as indicated by the z-score. Supplementary **Table S5** lists all 377 variants and their statistics.

### PWAS significant genes are sex dependent

A strong indication for sex dependent variants from GWAS results (**Figure 4**) raised the possibility of reanalyzing the cohort of males and females by a gene-based methodology. Importantly, the same variants were available for the GWAS and PWAS association methodologies (i.e., all imputed 639,323 gene coding variants). **Figure 5** shows the significant signal for the separated studies on males and females. We analyzed the 70 significant genes by PWAS (q-0.05; Supplementary **Table S4**). **Figures 5A** shows that for males, almost all genes were identified with a higher statistical confidence that is associated with genes in the unified cohort (i.e., signal below the diagonal). This is not the case for females with statistical values of genes also above the diagonal (**Figures 5B)**. A direct comparison of gene set identified in females versus males confirmed that 22 of 23 shared genes are significant in females and only one significant gene was identified in males relative to females (**Figure 5C**). For the PWAS q-0.1 set, six of 33 listed genes are significant in males relative to females (**Figure 5D**). Supplementary **Table S6** lists significant genes according to sex.

**Figure 5.**
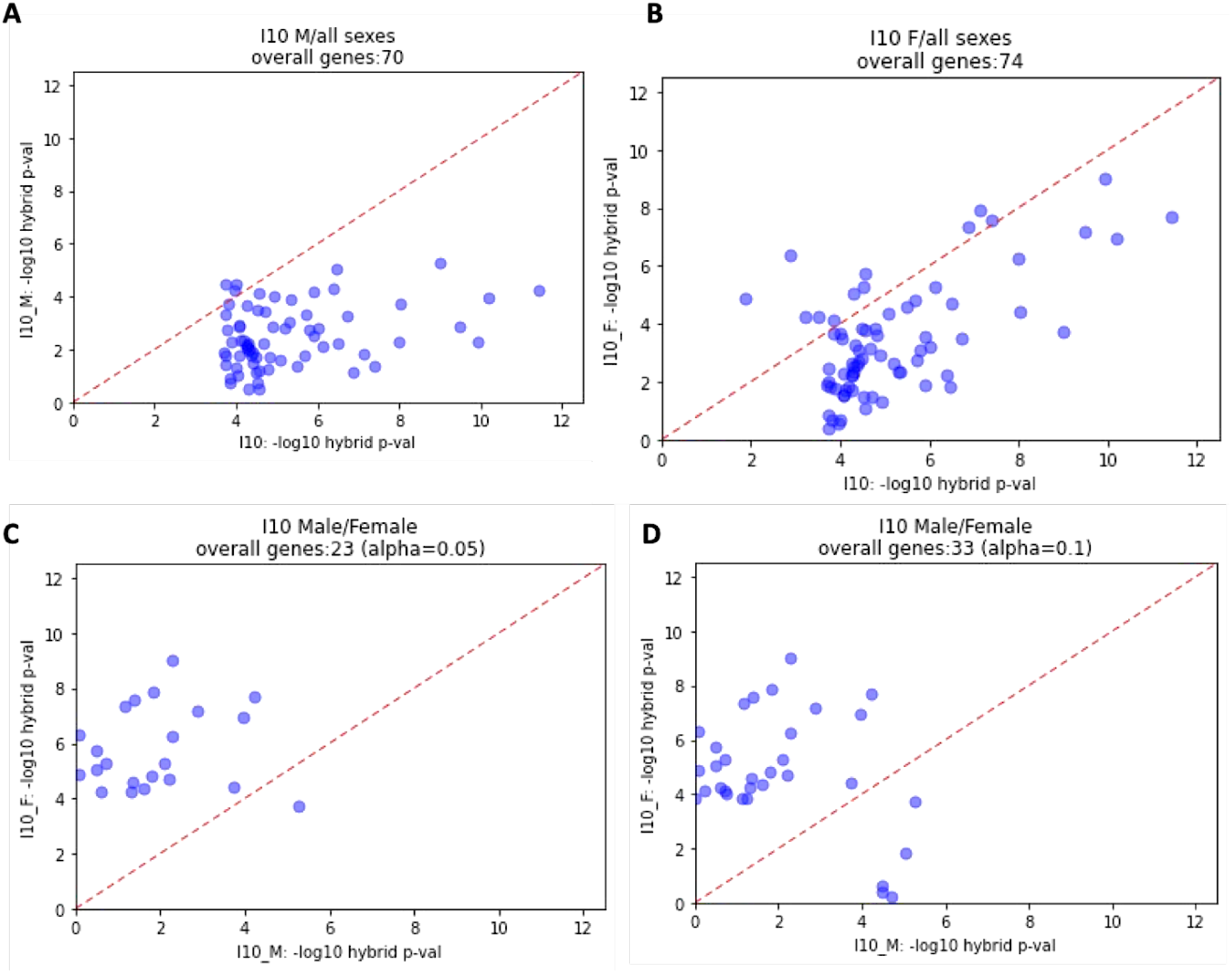
Sex partition for significant genes identified by PWAS. The 70 genes from the PWAS q-0.05 gene set were included in the analysis and converted to -log10(p-values). **(A)** Statistical significance for males vs. both sexes. **(B)** Statistical significance for females vs. both sexes. **(C)** Comparing significant genes in males vs. females in PWAS q-0.05. The analysis covers 23 shared genes. **(D)** Comparing significant genes in males vs. females in PWAS q-0.1. The analysis covers 33 shared genes. Only significant shared genes are shown based on a minimal significance of -log10(p-values)>4.0 for either males of females. Diagonal shows the expectation with no sex differences. M, males; F, females.

Venn diagrams that specify the overlap between PWAS results are shown in **Figure 6A** and **Figure 6B**. Remarkably, only SH2B3 overlaps between PWAS results for females, males, and both sexes. The significant genes of the female specific genes set (**Figure 6C**) and male specific genes set (**Figure 6D**) are sorted according to the statistical significance. While there were 28 significant PWAS (q-0.1) genes in females (22 included in the PWAS q-0.05 set), only six genes were indicated as significant for males, with only two genes included in the PWAS (q-0.05) set (Supplementary **Table S6**). We show that the PWAS-based signal is mostly restricted to females, in agreement with GWAS results (**Figure 4**). We concluded that females carry most of the genetic signal that dominates hypertension.

**Figure 6.**
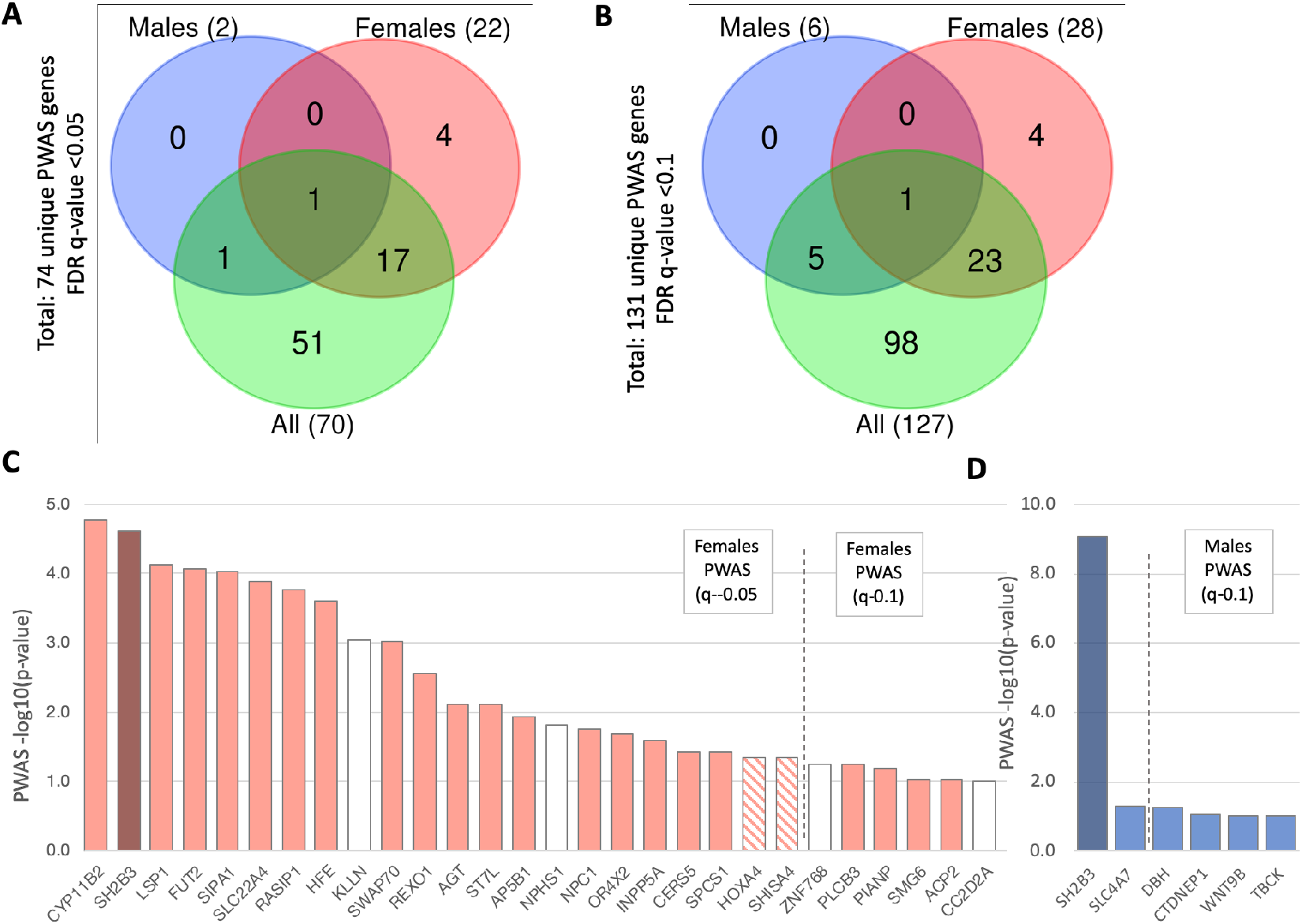
Sex dependent genetics of hypertension. Venn diagram for the overlap of the **(A)** PWAS (q-0.05) genes list, and **(B**) PWAS (q-0.1) genes list. **(C)** Ranked list of 28 genes significant in females for the PWAS (q-0.1; total 127), 22 of which are included in the PWAS (q-0.05) genes set (out of a total of 70). Dark color marks the single shared gene identified in (A). White color marks the genes that are specific for females. Striped pattern marks genes not included in the set of female exclusive PWAS (q-0.05) but not in the set of PWAS (q-0.1). **(D)** Ranked list of six significant genes of males in the PWAS (q-0.05) set of two genes, and another four included in the PWAS (q-0.1) set. Dark colors mark the single shared gene identified in (A). Vertical dashed lines partition the results of the PWAS (q-0.05) and (q-0.1). Supplementary **Table S6** lists the PWAS significant genes by sex.

### Cellular immunity related genes are enriched in PWAS-based results from females

One of the strongest advantages of PWAS is its ability to infer biological and biochemical effects of gene alterations. We therefore tested the underlying biology for the female-significant genes. We observed that many of the genes were mentioned in the modulation of the immune response. Results from GWAS (Supplemental **Table S1**) identified a set of significant variants in the introns and coding region of SH2B3 (also called Lnk, lymphocyte adaptor protein). SH2B3 is a member of the SH2B adaptor protein family that is involved in signal transduction. Our results identified it as having the largest effect on reducing risk for hypertension with respect to other associated genes. **Table 2** lists the most significant (top 15) genes identified by PWAS as female-specific associations. Half of the listed genes are maximally or uniquely expressed in immune organs (e.g., spleen, lymph nodes). The most significant PWAS identified gene in females is CYP11B2 (**Figure 6C**), which encodes a steroid 11/18-beta-hydroxylase (aldosterone synthase) that functions to synthesize the mineralocorticoid aldosterone. Aldosterone is a major regulator of intravascular volume and blood pressure. Interestingly, the expression of CYP11B2 is restricted to the adrenal gland where the expression in female tissue are 26% higher when compared to males (based on GTEx database). Moreover, the position of CYP11B2 in Chr 8q24.3 (Supplementary **Figure S3**) resides within the gene duplicated uPAR-LY6 gene cluster, which is active in immune cell recognition and mostly in autoimmunity. It is likely that the female specific manifestation of the gene combined with the immunological state drives sex-specific differences. The HFE protein is similar to MHC class I-type and regulates iron function. Individuals with dysregulation of iron metabolism are at higher risk for hypertension [31]. Moreover, the HFE was previously associated with essential hypertension in an independent cohort of Finnish subjects [32].

**Table 2.** Top 15 most significant PWAS genes in females

| Symbol | Name | <sup>a</sup>Sig (Dom, Rec) | Immune Function | Expression (Max) | <sup>b</sup>FDR q-value |
| --- | --- | --- | --- | --- | --- |
| CYP11B2 | cytochrome P450 family 11 subfamily B member 2 | Dom | Innate immunity | Adrenal gland | 1.70e-05 |
| SH2B3 | SH2B adaptor protein 3 | Dom | T cells & Macrophages | Spleen | 2.46e-05 |
| LSP1 | lymphocyte specific protein 1 | Both | T cells & DC migration | Spleen | 7.55e-05 |
| FUT2 | fucosyltransferase 2 | Rec | Autoimmunity (IBS) | Salivary gland | 8.55e-05 |
| SIPA1 | signal-induced proliferation-associated 1 | Both | MSC & memory T cells (cancer) | Spleen | 9.24e-05 |
| SLC22A4 | solute carrier family 22 member 4 | Both | Inflammation | Bone marrow | 1.30e-04 |
| RASIP1 | Ras interacting protein 1 | Rec | N.A. | Lung | 1.73e-04 |
| HFE | homeostatic iron regulator | Dom | Innate immunity, MHC-1 | Liver | 2.49e-04 |
| KLLN* | killin, p53 regulated DNA replication inhibitor | Rec | N.A. | Skeletal muscle | 8.93e-04 |
| SWAP70 | switching B cell complex subunit SWAP70 | Both | Lymphoid tissues, B cells | Lymph node | 9.75e-04 |
| REXO1 | RNA exonuclease 1 homolog | Dom | N.A. | Ubiquitous | 2.78e-03 |
| AGT | angiotensinogen | Both | Inflammation | Liver | 7.60Ee03 |
| ST7L | suppression of tumorigenicity 7 like | Rec | N.A. | Testis, Cervix | 7.61e-03 |
| AP5B1 | adaptor related protein complex 5 beta 1 subunit | Rec | Autoimmunity | Whole blood, Bone marrow | 1.16e-02 |
| NPHS1* | adhesion molecule, nephrin | Dom | N.A. | Kidney | 1.56e-02 |

<sup>a</sup>Sig, Significant for heritability by dominant, recessive of both; <sup>b</sup>FDR-q-value of the combined mode for females only. \* Significant only in females.

A set of 6 of the 28 genes are marked as exclusive to females (i.e., not included in the gene set for “all”; Figure 6C). Among the most significant PWAS genes (Table 2) is KLLN (Killin, P53 Regulated DNA Replication Inhibitor), which was studied in the context of cell division and cancer development, and it is not directly associated with the immune system. The other gene is NPHS1, which is mostly expressed in the kidney, and was previously implicated in sex-related differences in kidney aging [33]. While discovery of NPHS1 in females by PWAS was based on variants in the coding region, other intronic, synonymous, and upstream variants have been validated as pathogenic variants of NPHS1 and a cause of steroid-sensitive nephrotic syndrome (SSNS) [34]. Table 2 also shows that a third of the PWAS female significant genes are characterized by their recessive inheritance mode, and another third of the genes are significant in both inheritance models (recessive and dominant).

**Table 2** identified gene as associated with autoimmunity. Inspecting many GWAS studies that identified this gene with a strong association are signified with skin-related pathologies. Recently, it was shown that gene variants are causal for Atopic dermatitis in the Japanese [35] and Finnish populations (FINNGEN, freeze 5). Other cohorts identified the gene to be associated with psoriasis, asthma, hay fever or eczema. All these phenotypes are indicative of unbalanced cellular immune modulation. Inspecting the expression in whole blood, show a 30% elevation in expression in females relative to males (63.66 and 48.8 normalized TPM, respectively).

### Gene-based polygenic risk estimates for hypertension

For the goal of genetic-based personalized medicine PRS were calculated for hundreds of UKB phenotypes, including self-reported (non-cancer) hypertension (phenotype: HC215; with 83,727 cases of hypertension; Supplementary **Figure S4**) [36]. When hypertension PRS results are converted to prevalence by age (for 65 years), the prevalence is only 10% for the bottom 10% of the cohort, but 25.6% and 35.3% for the top 10% and 1%, respectively (Supplementary **Figure S5**). We argue that despite a clear genetic signal incorporated into the PRS for hypertension [36], interpretation or mechanistic understanding could not be reached. We used the PWAS identified gene lists to create gene-specific PRS-like statistics according to dominant and recessive considerations, as well as according to sex, and for the entire cohort. We examined the entire distribution of PWAS effect scores by considering all possible cutoffs for the relevant cohort, i.e., all subsets with effect scores that are either below or above a certain threshold. We describe the relevant sub-populations by percentiles (e.g., “bottom 10%” refers to the 10% of the cohort with the lowest PWAS scores) [26].

Figure 7 illustrates the PRS by a gene-specific hypertension risk for SH2B3 (SH2B adaptor protein 3; q-value = 9.9e-18) and CERS5 (Ceramide synthase 5; q-value = 2.2e-05). In the case of SH2B3, we observed that among the 434 individuals with the lowest dominant effect scores (comprising 0.12% of the cohort), 31.1% of that group had hypertension, compared to only 26.9 % in the rest of the cohort (leading to a quite modest observed risk-ratio of 1.16). We conclude that individuals with functional damage to the SH2B3 coding gene have a reduced risk of hypertension. In the recessive mode, only four individuals with the lowest recessive effect score (comprising 0.001% of the cohort), had hypertension (100%) relative to 26.9% in the rest of the sample, thus showing a risk-ratio of 3.7. CERS5 catalyzes the transfer of the acyl chain from acyl-CoA to a sphingoid base. As opposed to SH2B3, the dominant effect is maximal for the highest 13% effect scores (10,451 people), with 27.7% associated with hypertension. The perveance of the other 87% of the population is slightly lower at 26.8%. While the ratio difference is very small (1.03), there are over 10,000 people in this group. The recessive view is even more significant, with 28,792 people (39% of the tested set) with hypertension having 26.4% versus 27.3% with the rest of the population studied. These results suggest that CERS5 function increases the risk for many people, making it an attractive therapeutic target.

**Figure 7.**
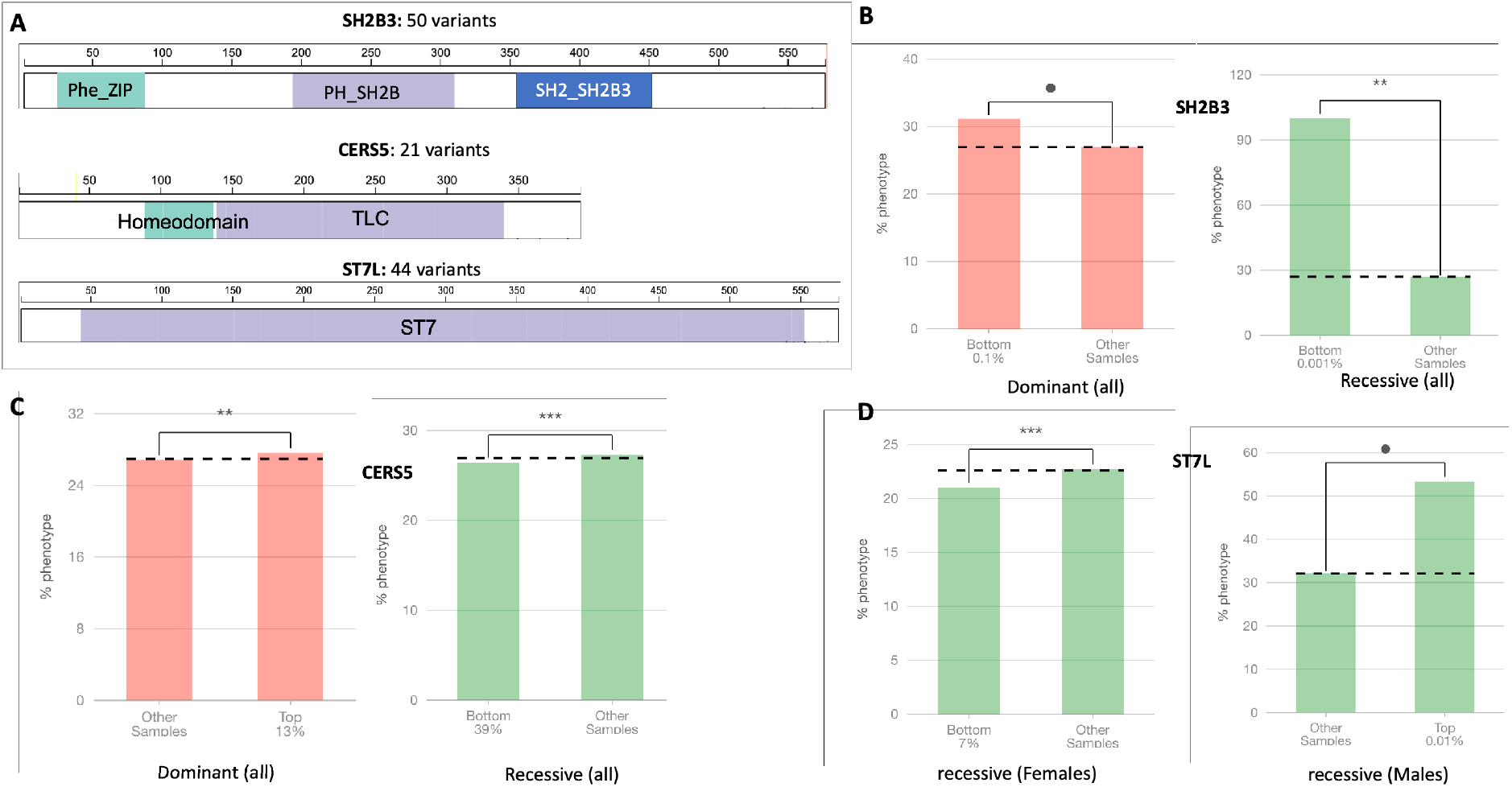
Dominant and recessive PWAS gene effects in PRS-like statistics. **(A)** Schematic view of SH2B3, CERS5 and ST7L genes along with their domains (labelled as colored rectangles). The length of the protein is indicated in amino acids, as well as the number of variants used to calculate the PWAS gene effect. (**B**) Dominant effect scores (red) captures the likelihood of an individual to have at least one damaged copy of the gene, whereas recessive effect scores (green) aim to capture the likelihood of at least two damaged copies. In PWAS effect scores, lower scores reflect individuals with greater gene damage. The plots show the most significant partitioning of the individuals (case-control status, selecting the cutoff with the lowest p-value by Fisher’s exact test). Note that each plot shows a different partitioning of the cohort (based on the relevant distribution of the effect scores). For example, the dominant effect scores ‘Bottom 0.1%’ for the dominant SH2B3 mode assigned to individuals that are the most likely to have at least one damaged copy of the gene. The prevalence of hypertension in the population is shown by the dashed horizontal baseline line across the entire cohort per gene. The significance of the results is indicated by asterisks. **(C)** Dominant and recessive effect scores for of CERS5 gene as in (B). (**D**) Recessive effect scores for ST7L genes for females and males. The partition for females for the bottom 7% of the cohorts suggest highly significant (***) for a reduced risk for hypertension. For males, the most informative PRS covers only 0.01% of the cohort, with a risk that was over 1.6-fold relative to the population.

**Figure 7D** demonstrates that the PRS-like analysis is valid for each of the sexes. While in females the recessive model shows a reduced risk of hypertension, in males a very small group (0.01%) exhibits a risk that is 1.6-fold relative to the rest of the population. Importantly, for the recessive model, I10 diagnosed in the entire population is 26.6% on average. It is 32.1% for males and 22.5% among women (21.1% for the 7% with affected gene, 2121 women), supporting its small but significant risk effect for ST7L gene in females.

## DISCUSSION

Hypertension is a complex physiological disease that involves the functions of the heart, the kidney, and the vascular system. In this study, we focused on essential (primary) hypertension diagnosis in order to test its underlying genetics in a large population. We used the UKB, which compiles information from 500k participants collected from 2006 to 2010. The analysis is limited to European ancestry (Caucasians) with unrelated individuals to minimize the genetic variations across ethnicities [37]. In addition, in most of the analyses, we included a large number of covariates (see Materials and Methods) to account for population structure and numerous technical biases. The SNP heritability estimate of 23.83% (p-value = 4.24e-116) was reported for UKB self-reported (non-cancer) hypertension. Hypertension heritability estimated that 31% of the BP variance is explained from genetics (ranging from 17-52% for systolic BP and 19-41% for diastolic BP, respectively) [37].

In this study, we used both GWAS and PWAS association methodologies and extensively compared them [25, 26]. We showed that PWAS as a gene-based method can detect non-additive effects **(Figure 3)**. PWAS assigns to each gene a dominant and a recessive effect score based on the genetic information available for that gene’s coding region. In the case of hypertension, about 18% of the PWAS-significant genes contain a recessive component that cannot be detected using variant-based methods. Many of the PWAS discovered genes with significant recessive effects, had not been previously identified by GWAS (**Figure 2**).

The majority of GWAS reports on variants found in intergenic and intronic regions, limiting their interpretability [15 52]. Identifying a gene from the variant(s) in GWAS necessitates additional evidence such as physical distances between the variants and the transcript start site, the location of regulatory regions (e.g., enhancers, chromatin), and other factors [38]. In fact, with over 1100 loci with leading SNPs, the transcriptome-wide summary statistics-based Mendelian randomization (TWMR) method [39] showed that 71% of the genes closest to that SNP do not show significant association with the tested phenotype. In contrast, PWAS assigns a gene by only considering variants (SNP markers and imputed) within the coding region, which allows direct functional interpretability. PWAS results are orthogonal to other association studies which rely on gene regulation [40].

The large scale of UKB (and other biobanks) allows an exploration of the genetics of sex differences. The majority of UKB phenotypes studied failed to show a genetic signal difference between males and females [41]. Sex differences in complex diseases were usually limited to anthropometric measurements (e.g., height, body fat distribution) and some biomarkers [42, 43]. Sex differences in hypertension was proposed to involve sex hormones. For example, when estrogen levels fall at menopause, the rates of hypertension increase. Accumulating data show that Ischaemic heart disease (IHD) exhibits a strong phenotypic sex difference, including atypical symptoms of IHD that are more common in women [44]. Moreover, estrogen acts on endothelial vasodilation primarily via inhibition of RAS (renin-angiotensin system) activity, causing a reduction in oxidative stress and inflammation [45]. Several genes were proposed to mediate the sex differences including ESR1 and ESR2 [46]. Still, the role of estrogen and gene function in vascular homeostasis is not well understood. We found that GWAS performed only on variants that are included in coding genes shows significant sex differences (**Figure 4C**) with a set of 106 variants in which the difference between the sexes (Δz) is large enough (defined by Δz >1e04). Interestingly, we identified a locus of Chr4 (position ∼107M) with a strong sex difference (**Figure 5B**). This locus was previously associated with hypertension and BP phenotypes (Supplementary **Figure S6**, top). The affected gene is TBCK (TBC1 domain containing kinase), which was also identified by PWAS (FDR p-value = 0.0486) and was also shown to be significant in males (**Figure 6D**). We propose that the combination of GWAS and PWAS for coding genes is a powerful approach to better assign variants to candidate causal genes.

The PRS-like analysis for genes (**Figure 7**) can be used to rationally use drugs. The gene-drug manipulations are likely to highlight attractive targets. For example, under the recessive model, the CERS5 (Ceramide synthase 5) and ST7L genes in females (but not in males) show that 39% and 7% of the relevant cohorts are at a lower risk for hypertension than the rest of the population, respectively. CERS5 has previously been linked to hypertension and BP traits [47]. A direct impact of CERS5 on cardiac autophagy and subsequent hypertrophy of cardiomyocytes underscored the importance of gene-specificity when treating complex pathologies [48]. The rationale is to identify the most likely gene that can be used in drug development for causal genes and to direct them to the appropriate population (i.e., females, males, or all). In future studies we will expand the observation to other biobanks from other ethnicities [49].

Lastly, we question whether the observed sex differences support an immunological-related explanation. Most immune system branches (T cells, NK cells, complement, macrophages) are involved in hypertension, according to animal models and large-scale human cohorts [50]. Cellular immunity, which results in chronic inflammatory responses of major organs (e.g., vascular system, heart, kidney), has been proposed to be the cause of hypertension [50]. Experimental models of hypertension in rodents confirmed the critical role of cells from lymph nodes and thymus in genetically modified and pharmacologically treated animal models (e.g., SCID mice). Nevertheless, knowledge of the molecular mechanism connecting common SNPs in hypertensive patients is sporadic and incomplete [51, 52]. Based on the PWAS and GWAS results, SH2B3 was highlighted as a protective gene in both sexes. This gene is associated with the strongest statistical signal, irrespective of sex (**Figure 5**). The SH2B3 nonsynonymous SNP (rs3184504) has been associated with numerous diagnoses such as ischemic stroke, hypertension and numerous autoimmune diseases (e.g., celiac disease, type 1 diabetes, rheumatoid arthritis, multiple sclerosis, vitiligo). The gene is also associated with systolic and diastolic BP and LDL cholesterol. While there is no animal model for SH2B3, causality has been proposed through the inflammatory and cellular immune systems [53]. In the established rodent model for salt-sensitive hypertensive rat (Dahl SS) a mutation in SH2B3 was linked to the attenuation of hypertension and renal disease. Importantly, it was shown that such attenuation in hypertension was carried out through SH2B3-mutated bone marrow cells, establishing a causal role of SH2B3 in inflammatory signaling which underlies hypertension [54]. For example, associations with cardiovascular disease death in the Framingham Heart Study with long-term follow-up confirmed the role of SH2B3 protein damage. Specifically, a decrease in SH2B3 expression led to higher levels of circulating B2M in the plasma. The SH2B3-B2M axis is a plausible mechanism for hypertension [53]. SH2B3 gene was also identified in the context of hypertension in the East Asian population [55].

Several genes were associated with the genetic effects only in females (**Table 2**) with HFE (homeostatic iron regulator) that may provide a link between the immune state (including autoimmune diseases) and iron metabolism [56]. The difference in the activation of the immune system in females is best reflected by the sex-bias of autoimmune diseases. FUT2, for example, was implicated in autoimmunity [57]. The association of such a mutation with Crohn’s disease was confirmed in diverse population origins [57] and showed an association with kidney disease in females [58]. Potential involvement of T cells in the development of hypertension was suggested using a transgenic male mouse model that lacks functional T and B cells. Adoptive transfer of T cells led to the restoration of the ability to manage BP in these mice [59]. Presumably, greater activation of the classical renin–angiotensin system (RAS) in males promotes the proinflammatory profile [60]. The increased expression of angiotensin receptor in females compared with males might affect the receptor’s role in controlling sodium excretion and renal function. Both estrogen and sex chromosome complementation are required for RAS function. In conclusion, we hypothesize that sex differences may not be a mere effect of hormones and sex chromosomal genetics but may expose yet unexplored evolutionary conserved biological mechanisms. Further studies are needed to better understand cellular immunity in hypertension. Such knowledge has potential medical implications for human health.

## Conclusions

Currently, interventions for hypertension are initiated later in life when BP values are already elevated and organ damage has already been initiated. Controlling hypertension is of great importance for preventing CVD, kidney failure, and other complications that reduce life expectancy. The SNP heritability of essential hypertension was shown to be high. Thus, genetics may be useful in assessing an individual’s response to anti-hypertensive drugs and minimizing their side-effects [61]. Our study showed sex-specific genetics for hypertension, a substantial contribution of the recessive component, and gene-specific personalized risk assessment. A molecular understanding of clinical hypertension in females proposes that the immune and inflammatory status are unexplored therapeutic targets. The incorporation of these considerations will lead to rational and personalized intervention in hypertension.

## Data Availability

All data produced in the present work are contained in the manuscript

## Data and code availability

The UK-Biobank (UKB) application ID 26664 (Linial lab). Ethical committee approval, The Hebrew University #13082019 from the University Committee for the Use of Human Subjects in research (dated 07/2002). PWAS is available through command-line interface as part of an open-source project (MIT license) at Brandes N. pwas. Github. 2020. https://github.com/nadavbra/pwas. Accessed 11 Apr 2020.

## Acknowledgments

We thank Dr. Nadav Brandes for the support in using the UKB parser, the PWAS algorithm and commenting on the initial draft. We thank Dr. Guy Kelman for his help in the implementation of PWAS. We thank Dr. Ronen Durst and Idit Gabay for sharing their views and commenting on the initial draft. We thank Amos Stern and Ido Blass from the Linial’s lab for useful discussions. We appreciate the constant support of the system team of the School of Computer Science at the Hebrew University.

## Abbreviations

BP: blood pressure
CAD: coronary artery disease
GWAS: genome-wide association study/studies
OR: Odds ratio
PRS: Polygenic risk score
RAS: Renin-angiotensin system
PWAS: Proteome-wide association study/studies
SNP: single nucleotide polymorphism
UKB: UK-Biobank

## Supplementary Tables

**Table S1:** Genetic-association normalized scores (ranges 0-1.0) from GWAS of hypertension (UKB self-reported). Total of 1362 genes have genetic associating evidence.

**Table S2:** A list of 205 SNPs with effective p-value <5e-08 from GWAS based on the UKB markers SNVs (804,069) associated with European origin.

**Table S3:** A list of 30 SNPs with from GWAS coding imputed genes with their effect size and statistics.

**Table S4:** A list of PWAS identified genes (total 127 genes) and their statistics.

**Table S5:** List of 377 variants with a minimal -log10>3.0 in only males or females.

**Table S6:** Listing the significant PWAS genes for females and males (28 and 6 genes, respectively).

**Figure S1.**
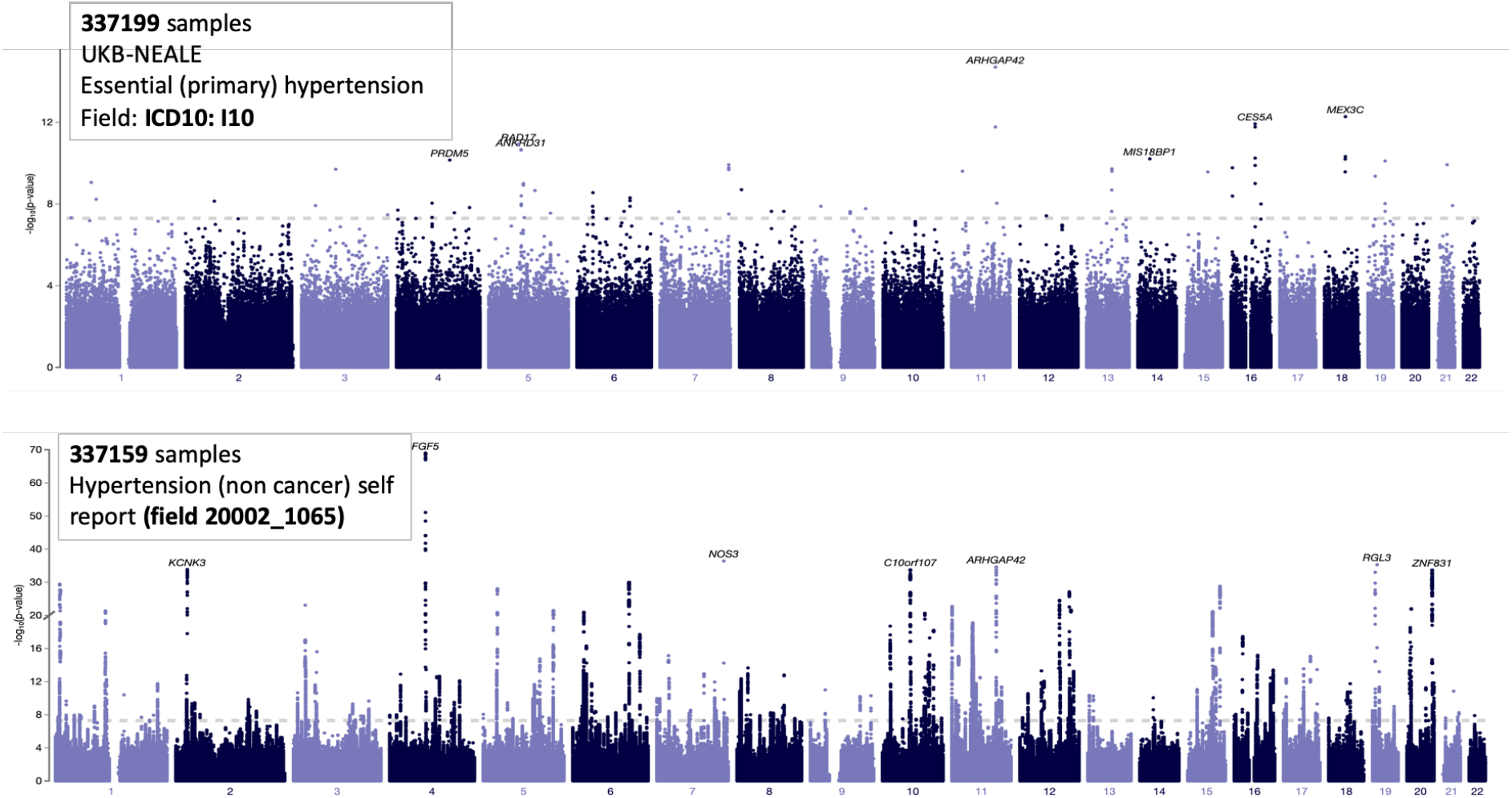
GWAS results for related phenotypes as reported by UKB for ICD-10 I10 (UKB main diagnosis, top) and hypertension (non-cancer) self-reported cohort (bottom). The difference is the finding and confidence is substantial (compare y-axis scale).

**Figure S2.**
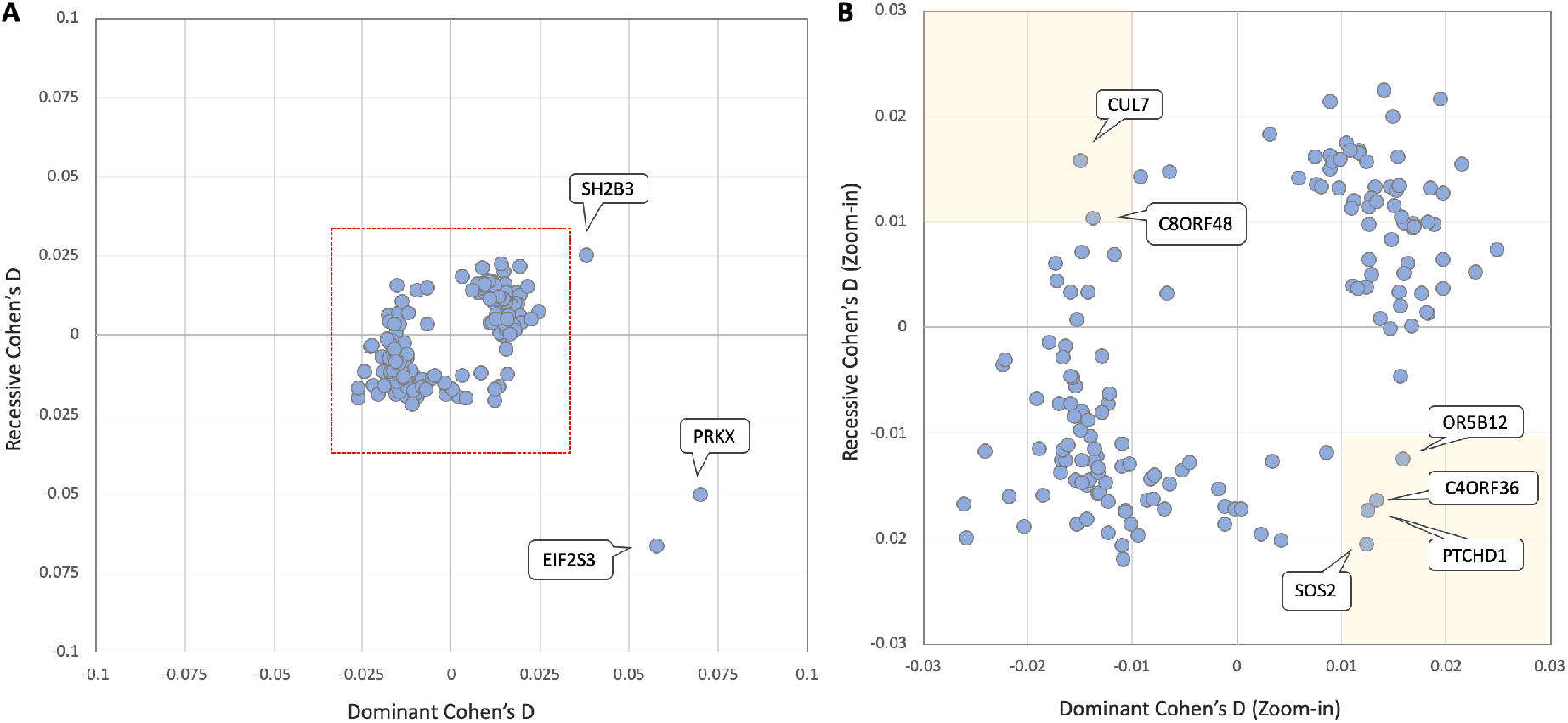
Effect size (Cohen’s d) for PWAS results for the recessive and dominant models. **(A)** The genes within the red frame are associated with Cohen’s d below an absolute value of 0.03 with positive values are associated with a reduced risk. The 3 genes with effect size >0.03 are marked. **(B)** Zoom-in for the genes with gene effects within value of -0.03 to 0.03. There are 6 genes that display apparently opposite directionality in Cohen’s d values (with effect size of at least >0.01 or <-0.01). The genes are labeled by their gene symbols. Supplementary **Table S4** lists all genes and their statistics.

**Figure S3.**
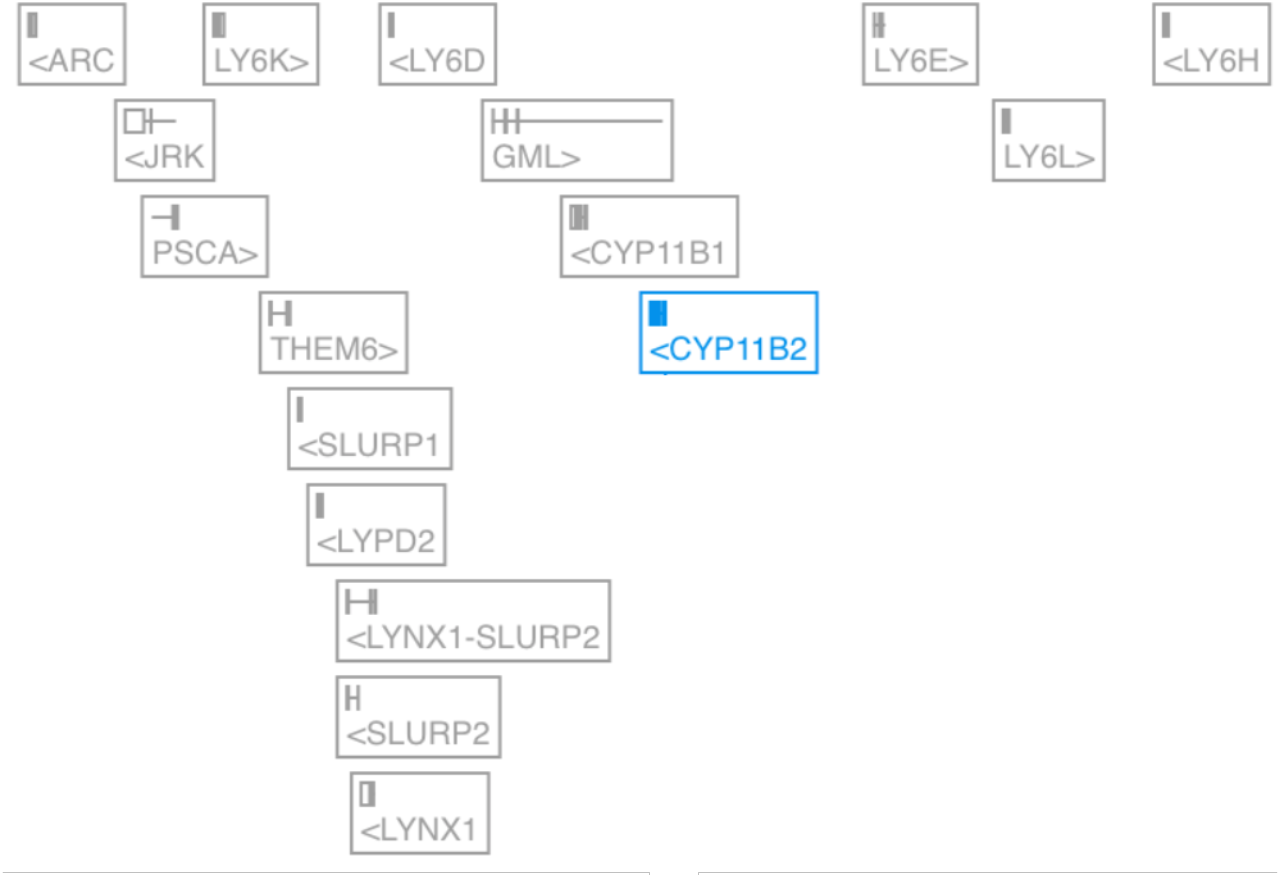
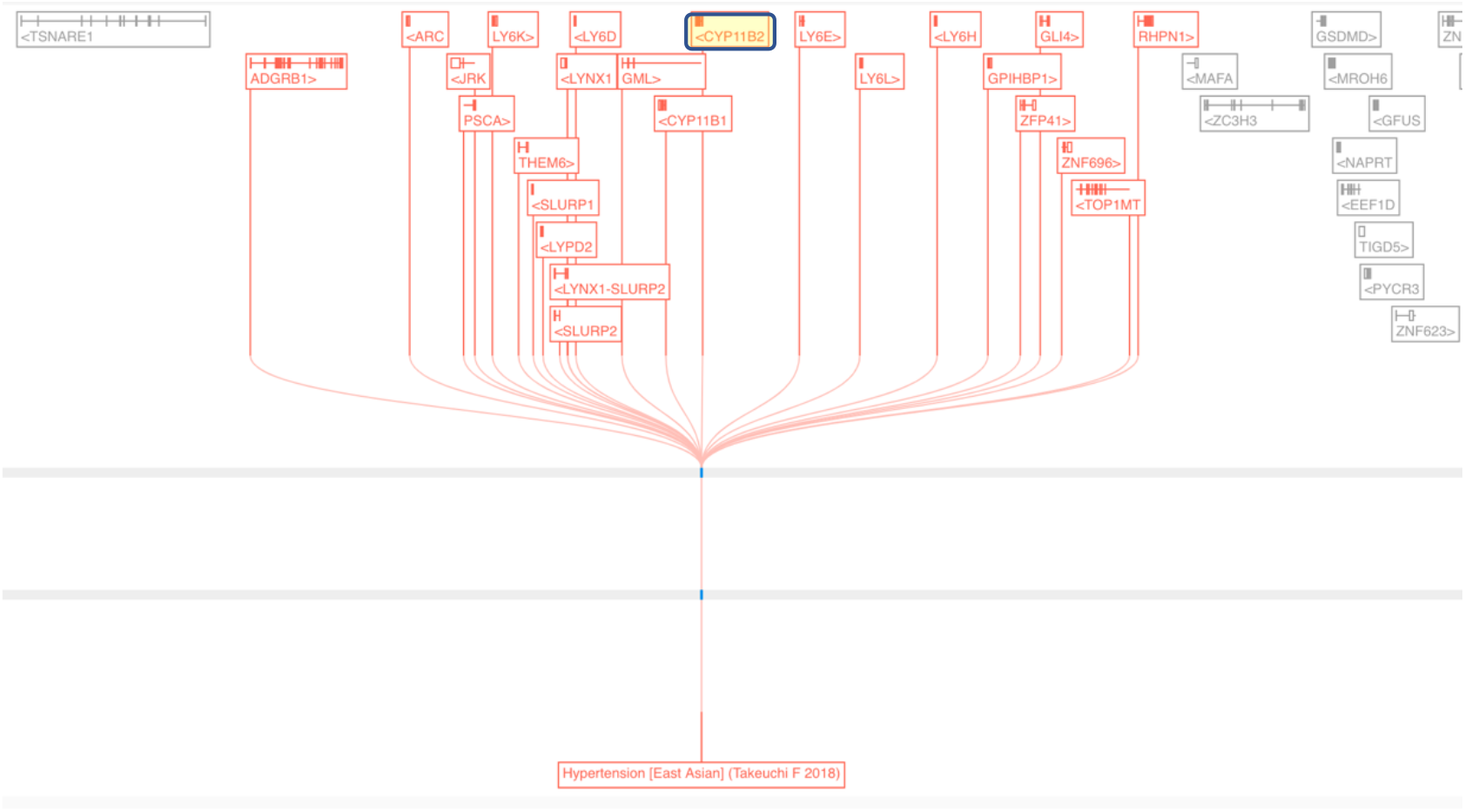
Genomic organization of CYP11B2 in Chr. 8q24.3. Top, the gene is a homolog is the neighboring gene CYP11B1 (93% identity). All the genes (in gray color) belong to uPAR-LY6 genes that dictate viral susceptibility and probably act in different aspects of innate immunity (e.g., SLURP2 regulates the pathophysiology of psoriasis [62]). Bottom, the locus of Chr8:142,910,559-142,917,843 from the Genetics OT platform assigned to the gene CYP11B2 (and a number of neighboring genes, shown in a red font). The variant rs12679242 within the intron of the CYP11B2 is exclusively associated with I10 diagnosis from East Asian population [55]. Notably, the minor allele frequency (MAF) of this variant is different across ethnic groups. For example, the population of East Asia and non-Finnish Europeans are marked with 0.298 and 0.477, respectively). The risk odds ratio associated with the leading SNP is 0.87, indicative of a protective variant.

**Figure S4.**
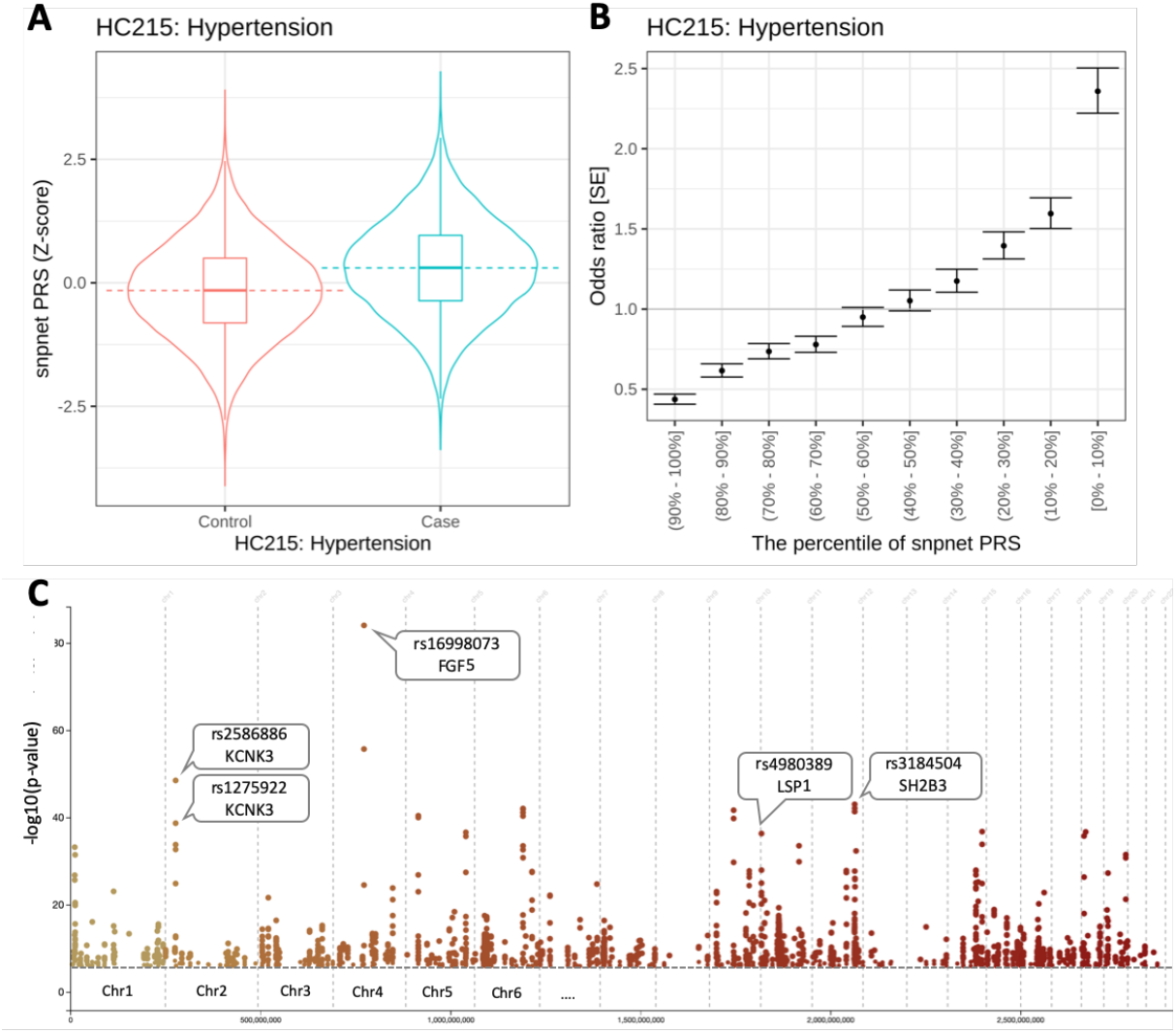
Analyzed multi-PRS from the snpnet PRS for UK population. **(A)** The PRS used 91,884 cases and 177,820 controls [36]. **(B)** The PRS is presented be deciles with OR (and standart error, SE). The analysis included genotypes and multiple covariates, and its predictive performance was assessed against a hold-out test set (22,882 and 44,543 for cases and controls, respectively) [63]. (**C**) Manhattan plots for hypertension from UKB. Several selected variants are labelled.

**Figure S5.**
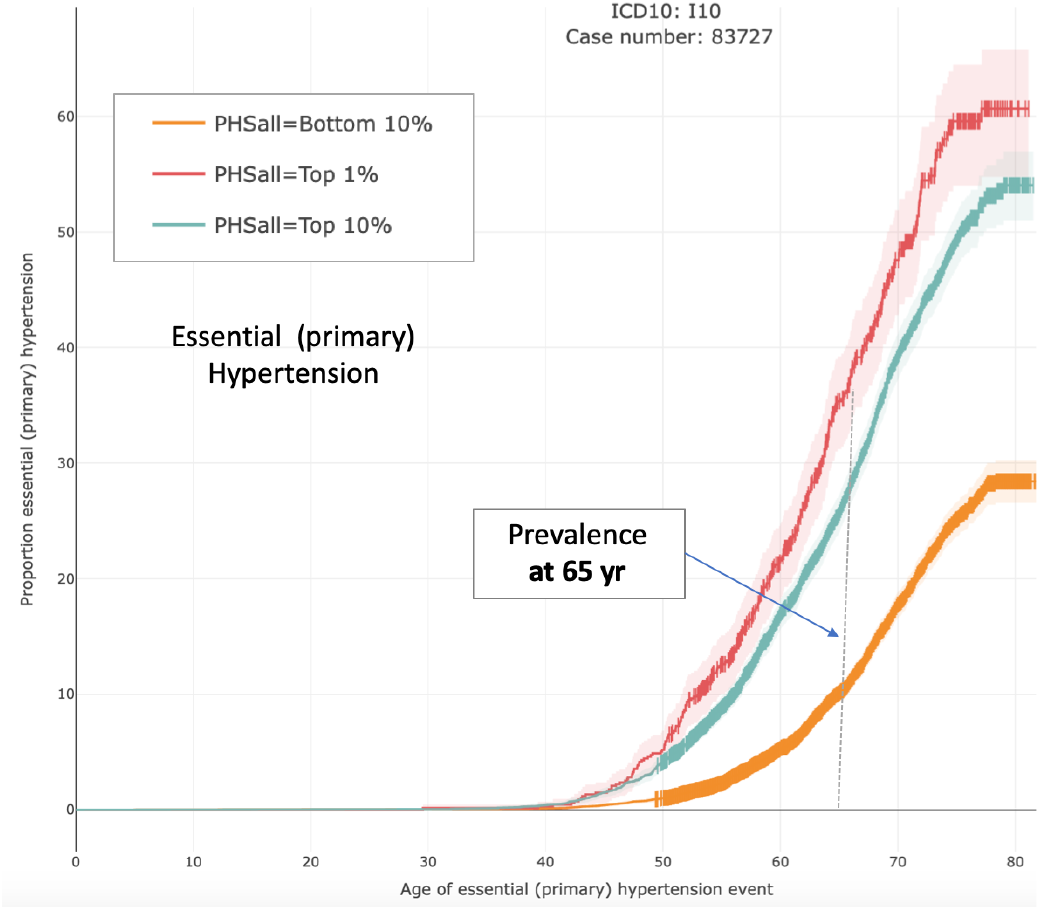
Age dependent PRS for ICD-10 of I10, partitioned by the lower 10%, upper 10% and upper 1%. Note the large difference in hypertension prevalence at age 65 (vertical dashed line).

**Figure S6.**
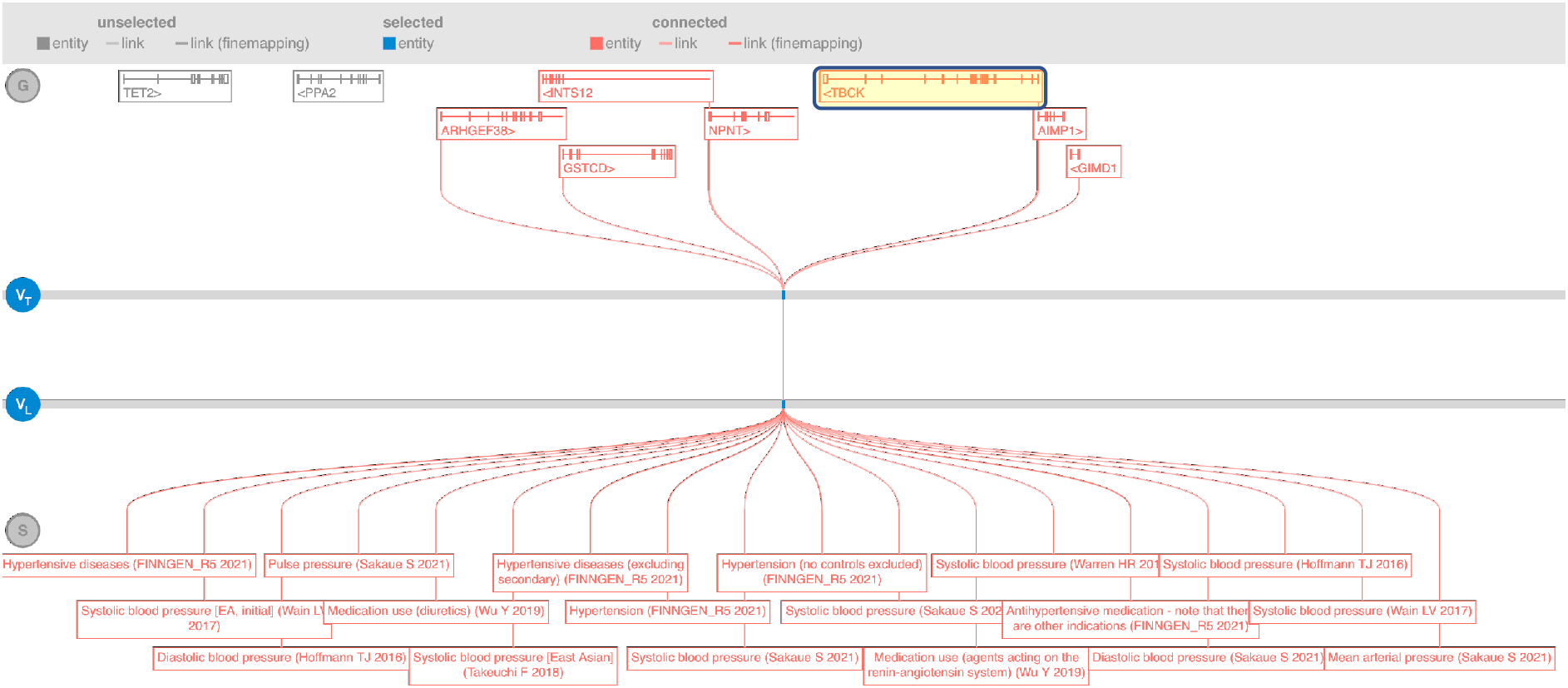
A locus Chr4:104,990,585-106,990,585 (Top) from the Genetics OT platform assigned to TBCK (and additional 6 neighboring genes). The leading variant that is associated with many studies of hypertension, including FINNGEN_R5_FG_Hypertension (2021), Systolic blood pressure study [64] and more (Top). The risk odds ratio associated with the leading SNP is 1.07.

